# Single-cell analysis identifies monocyte signatures of disease activity and clinical subtypes in Behçet’s disease

**DOI:** 10.64898/2025.12.19.25342559

**Authors:** Elio Carmona, Rabia Deniz, Cemal Bes, Haner Direskeneli, Ahmet Gül, Amr H Sawalha

## Abstract

**Objectives:** Behçet’s disease (BD) is a multisystem inflammatory disorder with diverse phenotypes and incompletely defined immune mechanisms. This study aimed to map immune dysregulation in BD at high resolution, comparing active versus remission states and identifying pathways linked to clinical phenotypes.

**Methods:** We performed single-cell RNA sequencing on 247,028 peripheral blood mononuclear cells (PBMCs) from 34 BD patients and 12 healthy controls. Transcriptomic profiling, differential gene expression, pathway enrichment analyses, and phenotype-stratified comparisons were used to delineate immune cell alterations associated with disease activity and clinical subtypes.

**Results:** All three monocyte subsets were markedly expanded in BD and demonstrated dominant IFN-γ–associated activation, robust heat-shock responses, and enhanced antigen-presentation programs. In active disease, monocytes exhibited pronounced type II interferon signatures, which reversed in remission alongside restoration of regulatory and metabolic pathways. Remission was instead characterized by increased expression of type I interferon-regulated genes, suggesting a potentially protective IFN-I-mediated effect, and by activation of SERPIN-associated programs linked to tissue stabilization. Clinical phenotype stratification revealed distinct monocyte signatures, with vascular BD enriched for heat-shock and stress-response pathways, and ocular BD defined by TNF/NF-κB-driven inflammation. Patients without organ involvement demonstrated an increased type I interferon gene signature, further supporting a potential protective role for type I interferon responses in BD.

**Conclusions:** This study provides a high-resolution immune atlas of BD, identifying monocyte-driven dysregulation as a central feature. Our findings map the immune heterogeneity of BD, identify activity- and phenotype-linked monocyte states, and suggest immune pathways suitable for targeted intervention.

## 1. Introduction

Behçet’s disease (BD) is a chronic, relapsing, multisystem inflammatory disorder characterized by recurrent oral and genital ulcers, uveitis, and systemic vasculitis that can affect vessels of all sizes and types[1, 2]. The disease exhibits substantial clinical heterogeneity, with vascular, ocular, and mucocutaneous involvement representing distinct phenotypic spectrums associated with variable severity and prognosis[2–4]. Although the etiology of BD remains elusive, converging evidence implicates an aberrant innate immune activation and dysregulated monocyte responses as central drivers of its pathogenesis[5–7].

Monocytes and their derivatives play a key role in bridging innate and adaptive immunity through cytokine secretion, antigen presentation, and crosstalk with endothelial and T cells[8, 9]. Aberrant production of proinflammatory mediators, such as TNF, IL-1β, IL-6, and type I interferons, has been reported in BD monocytes and peripheral blood mononuclear cells (PBMCs)[10, 11]. Yet, bulk transcriptomic and proteomic analyses have offered limited resolution to disentangle the contribution of individual monocyte subsets, which encompass classical (CD14⁺CD16⁻), intermediate (CD14⁺CD16⁺), and non-classical (CD14^dim^CD16⁺) populations with distinct immunoregulatory and tissue-repair functions[12, 13]. Understanding how these subsets are transcriptionally remodeled across disease activity states and clinical phenotypes in BD remain unaddressed.

Single-cell RNA sequencing (scRNA-seq) enables the dissection of immune cell heterogeneity and reveals cellular states driving complex diseases at unprecedented resolution[14]. Recent studies in immune-mediated conditions have uncovered disease-specific reprogramming of monocyte subsets, including interferon-responsive and cytotoxic phenotypes linked to disease flares and phenotype involvement[15–18]. However, a comprehensive single-cell map of BD immune cells, integrating both activity and clinical heterogeneity, is still lacking.

Herein, we performed single-cell transcriptomic profiling of PBMCs from patients with BD and healthy controls to define immune cell alterations associated with disease activity and phenotype. By integrating data from active and remission samples and across vascular, ocular, and mucocutaneous clinical forms, we provide an unbiased atlas of immune dysregulation in BD. Our analyses reveal transcriptional remodeling within the monocyte compartment, including the expansion of distinct monocyte subsets exhibiting chemotactic and interferon-responsive gene programs. These findings uncover shared and subset-specific molecular signatures underlying disease activity and organ involvement, offering new insight into the mechanisms driving immune heterogeneity in Behçet’s disease.

## 2. Materials and Methods

### Patient recruitment and clinical characteristics

Peripheral blood samples were collected from patients fulfilling the International Study Group (ISG, 1990) diagnostic criteria for Behçet’s disease (BD)[19]. Patient recruitment was conducted at Başakşehir Çam and Sakura City Hospital, Department of Rheumatology, Istanbul, Türkiye, and Istanbul University, Istanbul Faculty of Medicine, Division of Rheumatology, Istanbul, Türkiye. All participants provided written informed consent before sample collection. The study was approved by the Marmara University Clinical Research Ethics Committee (Approval No: 09.2021.52).

Active BD was defined as the presence of at least one new or recurrent clinical manifestation (oral or genital ulceration, ocular inflammation, or vascular involvement) within the last four weeks. All patients in the active BD group were newly diagnosed and treatment-naïve, having received no colchicine, corticosteroids, or immunosuppressive medications before sample collection. Remission BD was defined as the absence of any active clinical manifestation for at least four consecutive weeks following systemic therapy, with complete resolution of clinical signs and normalization of acute phase reactants. Clinical characteristics including age, sex, organ involvement, and disease status, were recorded at the time of sample collection (**Supplementary table 1**). Healthy controls were sex-matched healthy volunteers without a history of autoimmune or inflammatory diseases.

### PBMC isolation and single-cell RNA-seq protocol

Peripheral blood (20–30 mL) was collected into EDTA-coated tubes and processed within 2 hours of venipuncture. Peripheral blood mononuclear cells (PBMCs) were isolated by Ficoll-Paque density gradient centrifugation and washed twice with phosphate-buffered saline (PBS). No red blood cell lysis step was required. Cells were resuspended in freezing media (90% fetal bovine serum (FBS); 10% dimethyl sulfoxide (DMSO)), aliquoted at 5 × 10[ cells per vial, and frozen. Cells were thawed, washed and assessed for viability (>85% required). TotalSeq^TM^ B hashtag antibodies were then used for multiplexing samples in groups of 6[20]. Single-cell suspensions were loaded onto the 10x Genomics Chromium Controller for droplet encapsulation and library construction using the Chromium Next GEM Single Cell 3′ v4 HT kit, following the manufacturer’s protocol. Libraries were sequenced on an Illumina NovaSeq 6000 platform with a mean sequencing depth of over 11,000 reads per cell.

### Data preprocessing and quality control

Sequencing reads were aligned to the GRCh38 human reference genome using the Cell Ranger pipeline (v8.0.1, 10x Genomics). Resulting count matrices were imported into Seurat (v5.0)[21] for downstream analysis. Quality control filters included removal of cells with fewer than 400 detected genes, cells with >10% mitochondrial reads, and genes detected in fewer than 3 cells. Doublets were identified and removed using the DoubletFinder package[22]. Only high-quality cells were retained for further analysis.

### Clustering and cell type annotation

Filtered count matrices were normalized and variance-stabilized using SCTransform v2[23] with regression of mitochondrial content. Dimensionality reduction was performed using principal component analysis (PCA) accounting for >90% variation followed by Uniform Manifold Approximation and Projection (UMAP). Clusters in monocyte subsets were identified with the Louvain algorithm at a resolution of 0.4. Cell types were annotated based on canonical marker expression, supported by reference mapping to the Monaco Immune Database included as SingleR PBMC reference[24, 25]. Cell types represented by fewer than 200 cells were considered residual or low-confidence populations and excluded from downstream analysis to ensure adequate power and stability of statistical estimates.

### Differential gene expression and enrichment analyses

Differential gene expression (DGE) was assessed between BD and healthy controls, as well as between active and remission patient subsets and disease phenotypes, using Seurat’s FindMarkers function (Wilcoxon rank-sum test, Bonferroni/FDR-adjusted p < 0.05). Enrichment analyses of significant gene sets were performed using EnrichR[26] across Gene Ontology (GO:BP 2025) databases. For each differential expression contrast, upregulated and downregulated genes were analyzed separately to preserve directionality of regulation. Enriched terms with adjusted p-value < 0.05 were reported and ranked by the EnrichR Combined Score.

### Compositional analysis of immune cell subsets

Changes in immune cell subset proportions were analyzed using the sccomp package[27]. Unlike common proportion tests, sccomp provides probabilistic estimates of differential abundance that integrate uncertainty across subsets, yielding robust inference even in unbalanced designs. Relative frequencies of major PBMC subsets (T cells, B cells, NK cells, monocytes, dendritic cells) and subclusters were compared across BD patients and controls, and between disease activity states. Statistical significance was assessed using Bayesian proportion modeling.

### Patient and public involvement

Patients or the public were not involved in the design, conduct, or reporting of this research. However, we are committed to ensuring that the findings are shared in formats accessible to non-scientific audiences, including patients and the wider public.

## 3. Results

### Single-cell atlas of peripheral blood from patients with Behçet’s disease and healthy controls

We generated single-cell RNA sequencing profiles from peripheral blood mononuclear cells (PBMCs) of 34 patients with Behçet’s disease (BD) and 12 healthy controls (HC) to delineate immune cell heterogeneity and disease-related transcriptional changes (**Figure 1a**). After quality control filtering (removal of doublets and low-quality cells), a total of 227,735 high-quality cells were retained for downstream analyses. For active versus remission disease and longitudinal analysis, we also included 4 post-treatment samples of patients already included in the study, for a total of 247,028 cells. After clustering, cells were annotated into canonical PBMC populations using *SingleR* with *MonacoImmuneData[25]* as reference dataset and marker gene inspection. These included all major immune cell types (T cells, B cells, NK cells, monocytes, dendritic cells) that were broken down into 26 subcell types after removing residual populations (**Figure 1b, Supplementary figure 1**). The UMAP representation demonstrated clear separation of immune compartments, and integration after SCTransform normalization and scaling effectively minimized technical variation among sequencing pools (**Figure 1c**). The resulting dataset thus constitutes a comprehensive single-cell atlas of circulating immune cells in BD.

**Figure 1.**
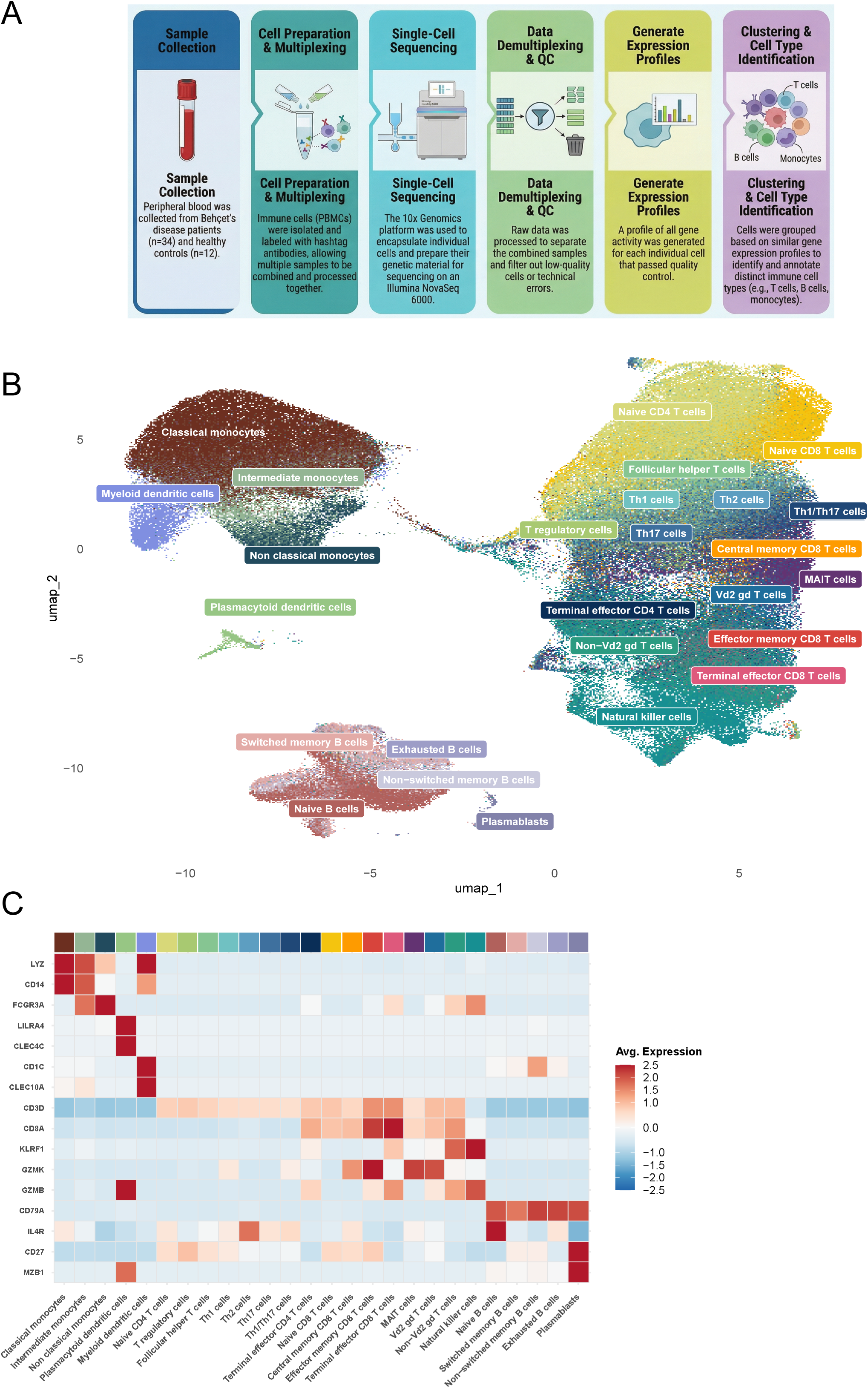
Study design and single-cell atlas of circulating immune cells in Behçet’s disease. (A) Overview of the study design. PBMCs were isolated from Behçet’s disease (BD) patients and healthy controls, spanning active and remission disease states and distinct clinical phenotypes, and profiled by single-cell RNA sequencing followed by demultiplexing, quality control, and integration. (B) UMAP projection of 247,028 high-quality PBMCs colored by annotated immune cell types, illustrating the overall cellular landscape and major compartments, including monocytes; plasmacytoid and myeloid dendritic cells; CD4⁺ and CD8⁺ T-cell subsets; natural killer (NK) cells; and B-cell subsets. (C) Heatmap of canonical marker genes demonstrating expression patterns that define each cluster and validate the annotation.

### Immune composition and transcriptional landscape distinguish Behçet’s disease monocytes from healthy controls

We first compared the cellular composition between BD patients and healthy controls. Monocytes were significantly expanded in BD compared to controls (effect = 0.486, 95% CrI = 0.169–0.8; FDR = 0.0098), corresponding to an approximately 1.57-fold increase in their predicted proportion relative to healthy controls (**Supplementary table 2, Supplementary figure 2a**). This imbalance in the monocyte compartment led us to further explore its cellular composition and transcriptional activity. Monocytes were subclustered and characterized according to established canonical markers, distinguishing between classical, intermediate, and non-classical subsets (**Figure 2a,b**).

**Figure 2.**
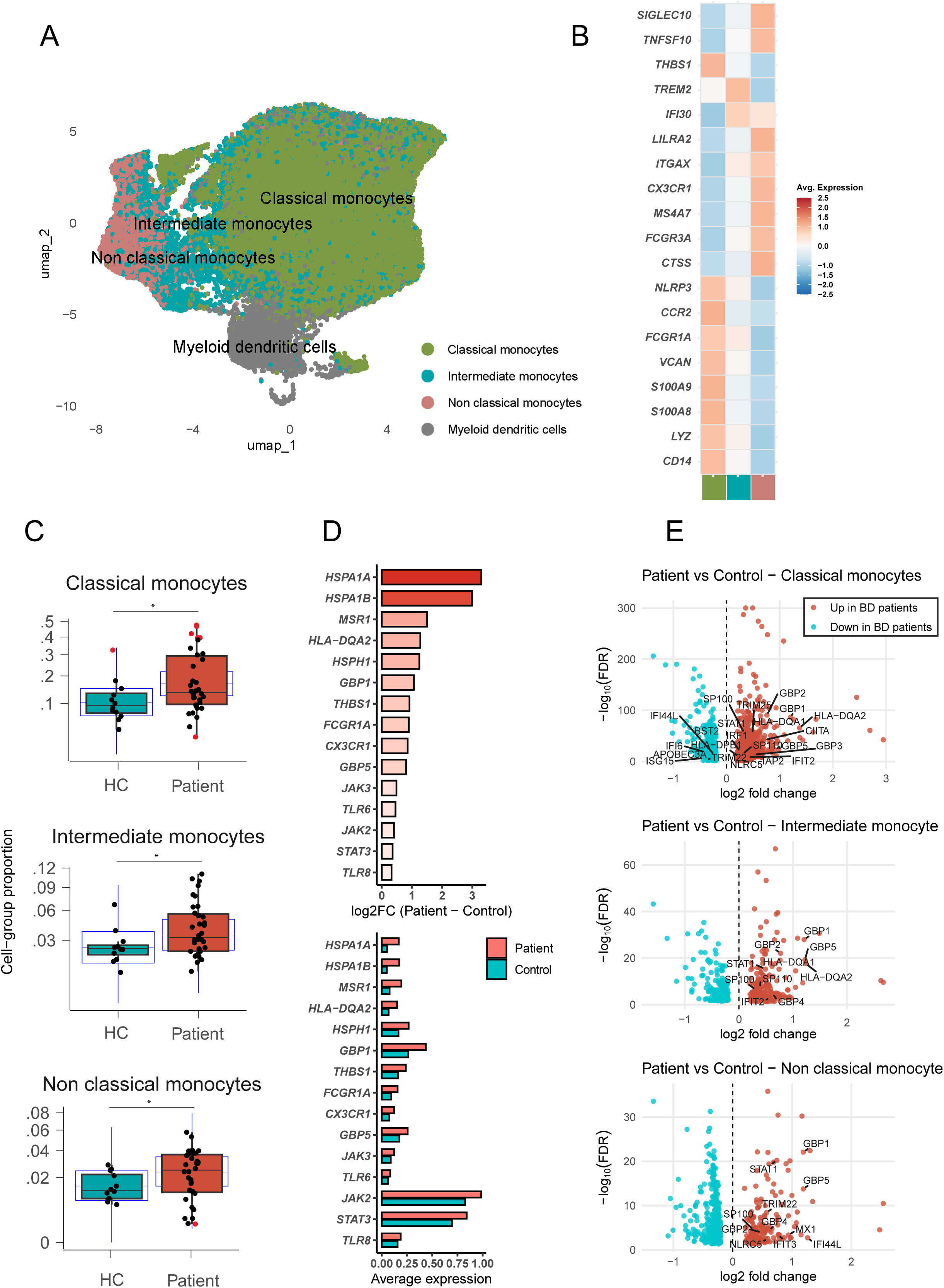
Monocyte subset heterogeneity and disease-associated transcriptional programs in Behçet’s disease. (A) UMAP projection of the monocyte compartment, delineating three transcriptionally distinct subsets: classical, intermediate, and non-classical monocytes. (B) Heatmap showing the top canonical marker genes defining each subset. (C) Cell-type composition analysis comparing the relative abundance of monocyte subsets between Behçet’s disease patients and healthy controls, illustrating significant expansion across subsets in BD. (*) indicate an FDR-adjusted *P*-value < 0.05. (D) Volcano plots of DEGs comparing Behçet’s disease patients versus healthy controls across monocyte subsets. Each point represents a gene, red indicating upregulation in BD and blue downregulation. The x-axis denotes the log₂ fold change, and the y-axis shows the –log₁₀(FDR). Only genes passing the significance threshold (FDR < 0.05) are shown. Canonical type I and type II interferon-stimulated genes (e.g., *GBP1*, *GBP2*, *GBP5*, *IFI27*, *IFI44L*) together with other ISG-related markers are labeled. (E) Barplot showing the top upregulated DEGs in Behçet’s disease monocytes compared with healthy controls. The upper panel displays log₂ fold-change values (patient–control). The lower panel shows the corresponding average expression levels in each group.

Compositional modeling revealed a significant expansion of all three monocyte subsets in BD compared with healthy controls. The most significant increase was observed in classical monocytes (1.64-fold, effect = 0.53, 95% CrI [0.22–0.83], FDR = 0.0018), followed by non-classical monocytes (1.49-fold, effect = 0.44, 95% CrI [0.08–0.80], FDR = 0.0116), and a milder yet significant increase in intermediate monocytes (1.35-fold, effect = 0.34, 95% CrI [–0.01–0.67], FDR = 0.0447) (**Figure 2c**). Beyond monocytes, myeloid dendritic cells, and non-Vd2 γδ T cells also displayed increased relative abundance, whereas naïve T cells, central memory CD8⁺ T cells and MAIT cells as well as non-switched memory B cells, exhibited decreased proportions in BD compared to healthy controls. These results highlight a coordinated expansion of the innate myeloid compartment alongside contraction of naïve T-cell pools, suggesting a systemic shift toward a pro-inflammatory immune landscape in BD (**Supplementary table 3, Supplementary figure 2b**).

To identify transcriptional alterations underlying the compositional changes in monocytes, we first examined differential gene expression across the monocyte compartment. BD monocytes displayed top upregulated genes related with inflammatory and interferon-responsive programs, including increased expression of canonical Interferon-regulated genes (e.g., *GBP1, GBP5, FCGR1A*), stress-response genes (*HSPA1A, HSPA1B, HSPH1)* and immune-activation markers such as (*MSR1, HLA-DQA2, THBS1, CX3CR1*) (**Figure 2e, Supplementary table 4**). Pathway enrichment analysis further revealed broad regulation of Interleukin-1 production, and response to type II interferon signaling, pinpointing the participation of genes like *JAK2/3, STAT3*, and TLR gene family members (e.g. *TLR6, TLR8*) (**Supplementary Table 5**).

We then analyzed each monocyte subset independently. Non-classical monocytes displayed transcriptional dysregulation characterized by upregulation of interferon-stimulated genes such as *GBP1–5, IFIT3, IFI44L, MX1,* or *STAT1* in BD compared to healthy controls (**Figure 2d, Supplementary table 6**). Pathway enrichment analysis revealed overrepresentation of type II interferon response, cytokine-mediated signaling, and antigen processing and presentation as predominant biological processes (**Supplementary table 7**).

Intermediate monocytes showed a similar pattern, with increased expression of *GBP1/5, MSR1, HLA-DQA1/2, SLAMF7*, and *GIMAP* family genes, together with enrichment of heat shock proteins (HSP), immune activation, and response to interferon and interleukin pathways (**Supplementary Tables 7,8**). In classical monocytes, which constitute the majority of monocytes in BD, there was marked upregulation of type II interferon-regulated genes in patients compared to controls, and downregulation of genes associated with the type I interferon signature (*IFI44L, IFI6, BST2, APOBEC3A, ISG15*) (**Figure 2d, Supplementary Tables 9**). Conversely, classical and intermediate subsets displayed reduced expression of *PHLDA2, DHRS3* and *TMEM* family genes which functions as regulators of cell stress signaling, metabolic homeostasis, and membrane trafficking (**Supplementary Tables 8, 9).** Notably, several transcripts were consistently suppressed across all monocyte subsets, including *G0S2, ABTB2, CXXC5,* and *HBEGF*.

We further dissected the myeloid compartment into eleven transcriptionally distinct clusters to characterize disease-associated programs (**Supplementary figure 3a**). Among these, cluster 7 displayed a prominent C1Q-high signature (*C1QA* avg. log_2_FC = 8.449; *C1QB* = 6.387; *C1QC* = 8.531; **Supplementary table 10**) along with higher *ISG15* and *ISG20*, which is consistent with a C1Q⁺ interferon-stimulated non-classical monocyte subset. This population, previously described in BD, represents a pro-inflammatory monocyte phenotype enriched in complement and type I/II interferon–response genes, and has been associated with disease activity and heightened innate immune activation[28, 29]. However, in our dataset, this specific subset, was slightly but not significantly expanded in BD compared with healthy controls (**Supplementary table 11).**

### Transcriptional reprogramming of monocytes differentiates active from remission Behçet’s disease

We next investigated molecular features distinguishing active from remission BD. Composition analysis revealed that all three monocyte subsets were significantly increased in frequency in active BD compared with healthy controls. The largest effect was observed in classical monocytes (effect = 0.62, 95% CrI = 0.33–0.92; FDR = 0.0005), corresponding to an approximate 1.73-fold increase respect to healthy controls. Non-classical and intermediate monocytes were similarly expanded (non-classical: 1.61-fold, effect = 0.55, CrI = 0.18–0.94, FDR = 0.0042; intermediate: 1.38-fold, effect = 0.41, CrI = 0.06–0.78; FDR = 0.0231) (**Figure 3a; Supplementary table 12, Supplementary figure 4a**). In contrast, remission BD showed a significant reduction in all 3 monocyte subsets compared to active disease indicating restoration toward control-like composition following clinical improvement (**Figure 3a; Supplementary table 13, Supplementary figure 4b**).

**Figure 3.**
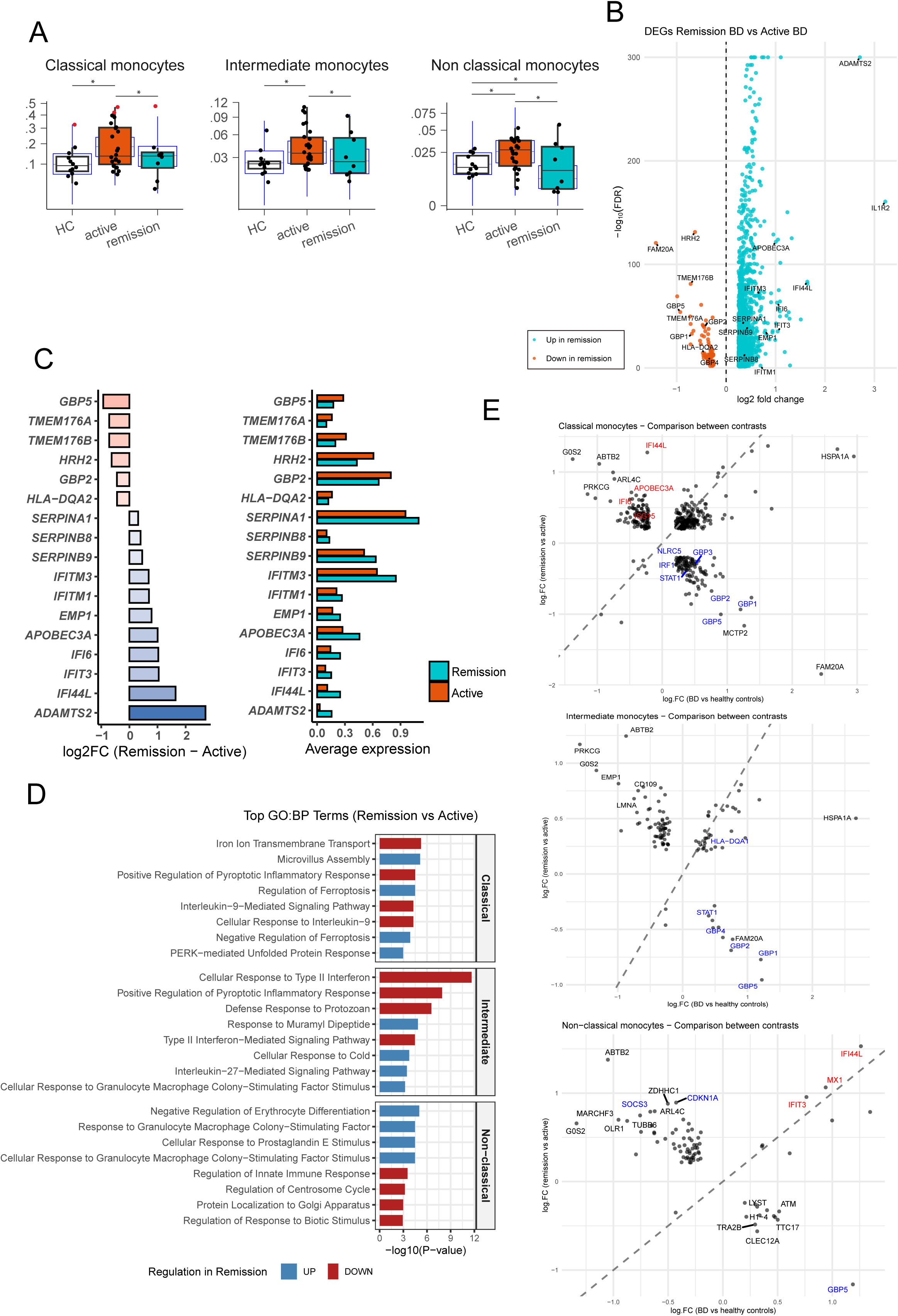
Monocyte subset remodeling across disease activity states in Behçet’s disease. (A) Compositional analysis comparing classical, intermediate, and non-classical monocyte proportions among active BD, remission BD, and healthy controls. Active BD is characterized by coordinated expansion of all three subsets, which partially normalize during remission.(*) indicate an FDR-adjusted *P*-value < 0.05. (B) Volcano plot of DEGs comparing active BD vs remission BD samples within all monocytes subsets. Each point represents a gene, with color indicating direction of expression change: orange denote upregulation in active BD, blue downregulation. All points are over the FDR > 0.05 threshold. (C) Differential expression summary of selected type I and II ISGs and SERPIN family members in remission versus active Behçet’s disease in total monocytes. Left panel presents the log₂ fold-change for each gene, with bar color and length indicating fold-change magnitude and direction (positive values reflecting higher expression in remission BD; negative values indicating higher expression in active BD). Right panel displays paired average expression values for active and remission samples. All genes showed FDR-adjusted *P*-value < 0.05. (D) Gene Ontology (GO:BP) analysis of the top 4 upregulated (red) and downregulated (blue) pathways compares remission versus active states across monocyte subsets. Significance is shown as −log_10_(P-value). (E) Quadrant plots depicting log₂ fold-change relationships between BD versus control (x-axis) and active versus remission (y-axis) comparisons for classical and non-classical monocytes. Genes in the lower-right quadrant (active^+^, BD^+^) represent disease-associated inflammatory programs, while those in the upper-left quadrant (remission^+^, BD^-^) indicate reversal genes restoring homeostatic or regulatory functions.

To define the transcriptional basis of these compositional changes, we compared gene expression profiles between remission and active BD samples. Remission monocytes displayed a broad shift away from the highly pro-inflammatory active disease state (**Figure 3b, Supplementary Table 14**). Type II interferon–associated genes, including GBP2, GBP5, TMEM176A/B, HLA-DQA2, and HRH2, were markedly reduced in remission compared with active disease, indicating contraction of the IFN-γ–driven inflammatory program. In contrast, canonical type I interferon–stimulated genes (IFITM1, IFIT1, IFIT3, IFITM3, IFI44L, IFI6, APOBEC3A, and EMP1) were more overexpressed in remission compared to active BD, suggesting a persistent and enhanced type I interferon imprint in monocytes during clinical improvement (**Figure 3b, 3c, Supplementary Tables 15–17**). Remission was also characterized by induction of a distinct set of recovery-associated transcripts. Genes including SERPINA1, SERPINB8, and SERPINB9 were coordinately increased in total monocytes during disease remission, reflecting activation of protease-inhibitory and cytoprotective programs that may contribute to tissue stabilization (**Figure 3b, 3c**). In classical monocytes, ADAMTS2 was upregulated, consistent with extracellular matrix remodeling and repair (**Supplementary Table 15**).

Consistent with these transcriptional changes, GO:BP pathway enrichment analyses revealed that classical and intermediate monocytes in active BD were dominated by type II interferon-driven inflammatory programs as well as cytokine-mediated pathways when compared to remission BD, underscoring a shared inflammatory core. In contrast, non-classical monocytes showed fewer and more heterogeneous pathways when comparing active versus remission BD, and lacked the prominent type II interferon and cytokine-response signatures observed in classical and intermediate subsets. Instead, remission non-classical monocytes were enriched for terms related to pharmacological responses, including cellular response to glucocorticoids. A similar enrichment of glucocorticoid response pathways was observed in intermediate monocytes in remission (**Figure 3d, Supplementary Table 18**).

To delineate how disease activity remodels monocyte states in Behçet’s disease (BD), we performed a reversal-gene analysis by jointly contrasting BD cases versus healthy controls and active versus remission BD for each monocyte subset. This two-dimensional framework identifies genes whose regulation across disease activity opposes their case–control behavior, highlighting transcriptional programs that invert with clinical improvement (**Figure 3e**). Classical monocytes displayed two major reversal patterns. Genes in the lower-right quadrant were upregulated in BD compared with healthy controls and downregulated in remission, representing inflammatory programs that reverse as disease activity resolves. Among the top features, *GBP1, GBP2, GBP3, GBP5* and *HLA-DQA2* exhibited strong reversal, along with *FAM20A* and *MCTP2*. Together with other ISGs like *STAT1, NLRC5* or *IRF1* suggest that type II interferon signaling, antigen presentation, and vesicular trafficking dominate the active inflammatory signature in classical monocytes that normalizes upon remission. On the other hand, upper-left quadrant genes were downregulated in BD but increased in remission, indicative of recovery of homeostatic and regulatory pathways. Top genes in this quadrant included *G0S2, ABTB2, PRKCG and ARL4C*, reflecting reactivation of metabolic control, cytoskeletal organization, and signaling balance lost during inflammation. Importantly, type I interferon-regulated genes, including *APOBEC3A, ISG15*, *IFI6* and *IFI44L*, also were downregulated in patients versus controls that became upregulated upon disease remission (**Figure 3e, Supplementary table 19**).

Non-classical monocytes revealed a similar but functionally distinct pattern. Genes in the lower right quadrant (*GBP5, CLEC12A, H1-4, ATM, TTC17, TRA2B, LYST*) were elevated in BD and downregulated in disease remission. In contrast, genes including *ABTB2, G0S2, SOCS3, ARL4C, ZDHHC1, MARCHF3,* and *OLR1*, were downregulated in BD but rebounded in remission, while the type I interferon markers *IFIT3, MX1* and *IFI44L* remained out of the reversal genes. These results reflect restoration of regulatory and metabolic programs in disease remission in non-classical monocytes, notably JAK-STAT negative feedback (*SOCS3*), lipid metabolism (*G0S2*), and cytoskeletal–adhesion reorganization (*ARL4C, TUBB6*) (**Figure 3e**, **Supplementary table 20**). Intermediate monocytes exhibited reversal signature in type II interferon-regulated genes similar to classical monocytes, along with rebound in genes including *G0S2, ABTB2, PRKCG,* and *HSPA1A* upon disease remission (**Figure 3e,Supplementary table 21**).

### Longitudinal paired analysis reveals partial normalization of interferon-driven inflammation after treatment

Among our BD cohort, four active patients were re-sampled in remission following therapy, enabling within-subject (paired) evaluation (**Supplementary tables 22-24**). For classical monocytes, some of the top DEGs up-regulated pre-treatment, *TMEM176A, TMEM176B, MCTP2,* and *FAM20A*, overlapped with those identified in the full active-versus-remission cohort. A similar trend was observed also in intermediate monocytes. GO:BP enrichment in classical monocytes emphasized bioenergetic/mitochondrial up-shift (e.g., *oxidative phosphorylation, electron transport chain, ATP synthesis*), with concurrent upregulation in type II interferon response and cytokine regulation pre-treatment compared to post-treatment in the same patients (**Supplementary table 25**). In intermediate monocytes, top pre-treatment GO:BP terms when compared to post-treatment samples reflect a strong IFN-γ-induced antigen-presentation signature, coupled with activation of monocyte/macrophage differentiation programs and concurrent regulatory pathways that modulate T-cell responses and inflammatory magnitude (**Supplementary table 25**). In non-classical monocytes, only two upregulated genes, *FMNL2* and *FCGR3A,* reached significance when comparing paired pre-treatment to post-treatment samples. These paired pre-/post-treatment samples showed moderate concordance with the full active-remission analysis across monocyte subsets, with consistent overall directionality (**Supplementary Figure 5**).

### Clinical phenotypes exhibit distinct immune signatures and intercellular communication patterns

To dissect clinical heterogeneity, we compared patients with vascular, ocular, and mucocutaneous disease-only subsets. We detected distinct distribution patterns among classical, intermediate, and non-classical monocytes (**Figure 4a, Supplementary table 26)**. In classical monocytes, patients with vascular involvement showed a significant increase in proportion compared with healthy controls (1.86-fold, effect = 0.59 [95% CrI 0.27–0.91], FDR = 0.0045), whereas mucocutaneous BD displayed a milder but still notable expansion (1.42-fold, effect = 0.39 [0.12–0.68], FDR = 0.0485). Ocular BD cases exhibited a trend toward higher abundance (1.14-fold, effect = 0.40 [–0.09–0.91]), though not statistically significant (FDR = 0.083). In contrast, intermediate monocytes showed no significant compositional shifts across phenotypes, with a similar expanded trend in mucocutaneous BD (1.48-fold, effect = 0.44 [–0.08–0.79], FDR = 0.063). Finally, non-classical monocytes demonstrated a marked increase in mucocutaneous BD (1.7-fold, effect = 0.56 [0.19–0.93], FDR = 0.023) but no significant change in vascular (1.18-fold, effect = 0.11 [–0.34–0.57], FDR = 0.34) or ocular BD (−1.77-fold, effect = –0.28 [–0.88–0.31], FDR = 0.20).

**Figure 4.**
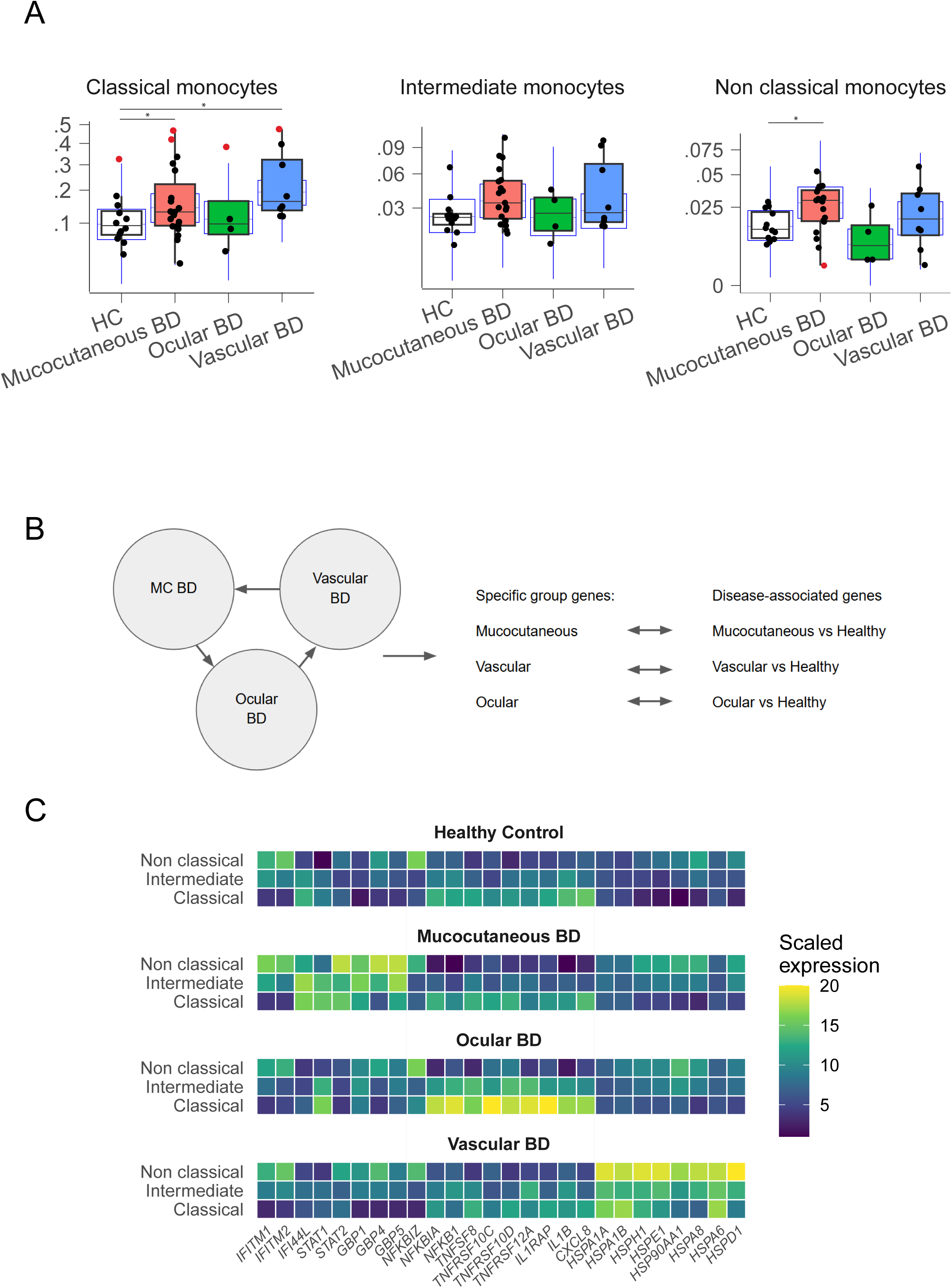
Monocyte compositional and transcriptional diversity across Behçet’s disease clinical phenotypes. (A) Compositional analysis comparing the relative abundance of classical, intermediate, and non-classical monocytes among Behçet’s disease phenotypes—mucocutaneous, ocular, and vascular—versus healthy controls. Distinct patterns of subset expansion are observed across phenotypes, with vascular BD showing the strongest enrichment in monocytes. (*) indicate an FDR-adjusted *P*-value < 0.05. (B) Schematic overview of the two-step filtering strategy used to define transcriptional signatures unique to each BD phenotype. In the first step, pairwise differential expression analyses were performed among BD phenotypes (mucocutaneous, vascular, ocular) to identify genes upregulated in each phenotype relative to the other two. In the second step, these upregulated genes were intersected with those upregulated in the same phenotype versus healthy controls, yielding phenotype-specific, disease-associated gene sets. (C) Tile plot illustrating phenotype-specific transcriptional programs within each monocyte subset. Interferon-stimulated genes (e.g., *IFI44*, *IFITM1/2*, *GBP1/4/5*), NF-κB and TNF signaling mediators (*NFKB1*, *TNFRSF10A/D*, *IL1B*), and stress-response chaperones (*HSPA1A/B*, *HSP90AA1*) reveal differential activation signatures across phenotypes, highlighting shared and subset-specific immune programs.

We next sought to identify phenotype-specific transcriptional programs across vascular, ocular, and mucocutaneous involvements. Pairwise differential expression analyses were performed within each cell type to pinpoint genes upregulated in one phenotype relative to the other two. To ensure these genes reflect features unique to that phenotype in disease, we then required that they also be upregulated in that phenotype compared with healthy controls. This two-step approach isolated a set of *phenotype-specific, disease-associated signatures* while excluding genes that are common to all BD cases (**Figure 4b**). Distinct inflammatory modules were observed. Mucocutaneous BD was characterized by broad upregulation of interferon-stimulated genes (*IFITM1, IFITM2, IFI44, GBP1, GBP4, GBP5, STAT1,* and *STAT2*) across classical, intermediate, and non-classical monocytes. In contrast, vascular BD, particularly in non-classical monocytes, showed strong enrichment for heat-shock and stress-response genes (*HSPA1A, HSPA1B, HSP90AA1, HSPH1, HSPD1*), consistent with heightened cellular stress and protein-folding demand under inflammatory or oxidative conditions. Meanwhile, ocular BD displayed prominent expression of TNF/NF-κB-associated cytokines and receptors, including *IL1B, CXCL8, NFKB1, NFKBIA, NFKBIZ,* and *TNFRSF10C/D* (**Figure 4c, Supplementary table 27**). These polarized transcriptional programs were most evident in monocytes but were also partly mirrored in other immune populations (**Supplementary table 27**), suggesting that phenotype-specific inflammatory signatures (interferon signaling in mucocutaneous BD, stress-response activation in vascular BD, and TNF/NF-κB–mediated inflammation in ocular BD) extend beyond the myeloid compartment and may underlie the divergent clinical manifestations of the disease.

## 4. Discussion

Our study provides a comprehensive single-cell view of circulating immunity in Behçet’s disease across activity states and clinical phenotypes. By profiling 247,028 PBMCs from 34 BD patients and 12 healthy controls, we delineate a robust atlas in which the monocyte compartment emerges as the principal locus of dysregulation. Compositionally, our data support coordinated expansion of monocyte subsets in BD, while transcriptionally we resolve subset-specific programs that differentiate active disease from remission and vary across vascular, ocular, and mucocutaneous phenotypes. Together, these data refine current models of BD pathogenesis by linking clinical heterogeneity to discrete myeloid activation states, and suggest that phenotype-specific targeting of innate pathways may better align with the underlying cellular biology.

Behçet’s disease is increasingly recognized as a monocyte/macrophage-driven vasculitis, with circulating monocytes showing M1-skewed activation, enhanced TLR expression, and elevated production of TNF-α, IL-6 and IL-1 family cytokines[7]. Previous studies have reported an imbalance of CD14/CD16-defined subsets characterized by expansion of intermediate monocytes and contraction of non-classical monocytes[30],a pattern interpreted as enhanced recruitment of CD16⁺ monocytes to inflamed tissues. In this context, our findings of an overall expansion of circulating monocytes in BD, driven by significant increases in all subsets compared with healthy controls, indicate that the entire compartment is numerically amplified in BD rather than a single subset being selectively depleted. Although partially divergent from previous reports, these findings still fit within a broader model where monocyte dysregulation and activation are key to BD pathogenesis. Additionally, the even stronger expansion of all three subsets in active disease and their partial normalization in remission mirror prior observations of monocyte frequencies declining with effective therapy[28].

Prior transcriptomic and single-cell studies have converged on a prominent type II interferon imprint in BD monocytes. Bulk and tissue-level analyses show that differentially expressed genes in BD are enriched for IFN-γ–response pathways, with upregulation of CXCR3 ligands (*CXCL9, CXCL10*) and guanylate-binding proteins (*GBP4, GBP5*)[29]. Single-cell profiling of PBMCs further identified an expanded CD14⁺ C1Q^hi monocyte population driven by IFN-γ–JAK–STAT signaling and reversible with tofacitinib, directly linking IFN-γ dependent monocyte programs to hyperinflammation in vivo[28]. In parallel, functional work has shown that BD monocytes display dysregulated post-transcriptional control of CXCL10, leading to exaggerated secretion of this IFN-γ–inducible chemokine and thereby amplifying Th1-cell recruitment upon IFN-γ stimulation[31]. Together, these studies support a model in which IFN-γ positions monocytes at the core of BD immunopathology, reprogramming them into antigen-presenting, chemokine-producing effectors that sustain Th1-skewed, vasculitic inflammation.

In this context, our data extend previous work by showing how this IFN-γ activation is extended to the broader monocyte compartment. As a whole, BD monocytes upregulated canonical type II interferon and inflammatory genes (e.g. *GBP1/5, JAK2/3, FCGR1A, HLA-DQA1/2, CX3CR1*) compared to HC, with enrichment of IL-1 production, and response to type II interferon, indicating that IFN-γ shapes both effector function (phagocytosis, Fc receptor signaling) and tissue-interacting properties (adhesion, chemotaxis). Across classical, intermediate, and non-classical subsets we observed consistent induction of IFN-γ–responsive modules, while classical monocytes showed broader type II interferon signature coupled to downregulation of some canonical type I ISGs. In line with prior data, our findings point towards a ubiquitous type II interferon signature across classical, intermediate, and non-classical monocytes that amplifies IL-1 and JAK–STAT–dependent inflammation and sustains the hyperinflammatory milieu characteristic of BD.

The role of type I interferon in BD however, appears to be more complex and context dependent. Exogenous IFN-α has well-established therapeutic efficacy in mucocutaneous and ocular BD, where it can dampen pathogenic Th17 responses and promote regulatory circuits[32–34]. In this sense, we observed that classical monocytes, which dominate the circulating monocyte pool, showed a clear skewing toward a type II interferon program in BD, with marked upregulation of IFN-γ–regulated genes and concomitant downregulation of canonical type I ISGs (*IFI44L, IFI6, BST2, APOBEC3A, ISG15)*. By contrast, non-classical and intermediate monocytes upregulated a more restricted set of ISG-family genes often labeled as type I responsive (such as *IFI44L, IFIT3, IFI16* in non-classicals and *IFI16, IFIT2* in intermediates) in BD versus controls. This pattern reflects that BD is not a type I interferonopathy in circulating monocytes, but rather an IFN-γ/IL-1–dominated activation state in classical monocytes with selective type I ISGs induction in CD16⁺ subsets (Intermediate and non-classical). Consistent with this idea, we identified a C1Q^hi/ISG15^hi monocyte cluster (**Supplementary table 10**) that closely matches the C1Q^hi and ISG15^+ monocytes described in previous studies where BD skin lesions show expansion of ISG15^hi monocytes with strong expression of ISG15 and other ISGs such as IFI44L together with C1Q genes, consistent with IFN-like stimulation within inflamed lesions[29]. A recent single-cell transcriptomic study further demonstrated mobilization of inflammatory C1Q^hi monocytes in BD blood and their accumulation as C1Q^+ and ISG15^+ monocytes within skin, underscoring a direct contribution of activated monocyte subsets to tissue inflammation[28].

Notably, many of the ISGs induced in non-classical and intermediate monocytes are shared targets of both type I and type II interferons and can also be driven by other inflammatory factors, particularly in patrolling cells engaged in antiviral and endothelial surveillance[35]. In this sense, an unexpected finding was the differential behavior of some type I versus type II interferon genes across disease activity. While IFN-γ–associated signatures (*GBP1/5, FCGR1A, CCL4, IL15RA, TMEM176A/B*) showed marked reduction in patients with remission BD, previously mentioned type I ISGs (*ABTB2, ISG15, IFI6, IFI44L, APOBEC3A*) remained elevated or were even more prominent in remission monocytes. This decoupling explains how current therapies may effectively suppress the most defined IFN-γ–driven inflammatory circuits while leaving behind a persistent type I interferon imprint.

When examined, clinical activity at subset level revealed that interferon modules likewise did not behave uniformly across disease states. In active BD, monocytes were dominated by a type II interferon driven program, with strong induction of GBP genes and antigen-presentation pathways. By contrast, in remission these GBP-associated, IFN-γ–linked signatures declined while canonical type I ISGs (e.g., *IFI44L, IFI6, IFIT3, IFITM1/2/3, APOBEC3A*) were consistently upregulated across all monocyte subsets and particularly in classical monocytes. Together with the observation that this canonical type I ISG module is selectively downregulated in BD versus controls in classical monocytes, the recovery of type I ISG expression in remission supports a model in which IFN-γ–skewed programs are pathogenic in BD, while a homeostatic type I interferon circuit may be relatively protective. This approach supports previous literature, where Th1/Th17 cytokines (IFN-γ, IL-12, IL-23, IL-17) are repeatedly implicated as pathogenic in BD[36]. The recovery of *APOBEC3A* in classical monocytes, a type I ISG recently shown to limit STAT1-dependent ISG expression, is consistent with re-establishment of a feedback-regulated type I interferon network in remission rather than ongoing uncontrolled interferon activation[37]. Similarly, *SOCS3*, a JAK–STAT feedback inhibitor[38], was decreased in intermediate and non-classical monocytes from BD patients yet selectively increased in remission in non-classicals, consistent with re-engagement of negative-feedback circuits that restrain IL-6–family and interferon signaling in this patrolling subset. Such SOCS3 induction in remission BD would be expected to limit residual type I and II IFN signals, potentially protecting against relapse of Th1/Th17-driven inflammation, while also raising the possibility that homeostatic type I IFN-mediated antiviral and endothelial-surveillance programs are attenuated in non-classical monocytes. Although exploratory, these signals raise the possibility that disrupted negative regulators contribute to the reshaped interferon landscape in BD and ask for a functional follow-up.

Based on our data, monocyte activation in active BD is dominated by a type II interferon– and IL-1–skewed program, marked by GBP and HLA upregulation, whereas canonical type I interferon–stimulated genes are relatively attenuated in this state and rebound in remission. Thus, this pattern supports therapeutic strategies like ustekinumab and JAK inhibitors that dampen IFN-γ/IL-12/23–JAK–STAT signaling[39, 40]. By contrast, the partial recovery of a canonical type I ISG module in remission, together with prior clinical experience showing efficacy of IFN-α in refractory Behçet uveitis[41], is consistent with the notion that type I interferon can exert net immunomodulatory rather than pathogenic effects in BD. Definitive conclusions regarding type I IFN agonism versus antagonism, however, will require dedicated mechanistic studies.

In addition, remission BD monocytes were marked as well by a serpin-enriched, tissue-preserving program. The observed increase in *SERPINB2*, *SERPINB9*, and *SERPINA1* expression among this compartment suggests that remission in Behçet’s disease involves more than simple suppression of inflammation. SERPINB2 (PAI-2) and SERPINA1 (PAI-1) modulate fibrinolytic activity and limit proteolytic tissue injury, whereas SERPINB9 (PI-9) protects cells from granzyme B-mediated apoptosis, conferring resistance to NK and CD8⁺ T-cell cytotoxicity[42]. This serpin-enriched signature implies a transition toward a cytoprotective and remodeling phenotype, supporting vascular repair and sustained immune homeostasis. Together with the downregulation of interferon-driven inflammatory genes, these findings highlight that effective therapy in Behçet’s disease could rebalance monocyte function from proinflammatory activation to tissue preservation and immune regulation.

By looking at phenotype distribution of cells, the expansion of circulating monocytes observed in BD patients compared with controls were largely driven by the mucocutaneous and vascular phenotypes, where classical and non-classical subsets respectively showed the most pronounced increases. In contrast, intermediate monocytes exhibited a more uniform distribution across phenotypes, suggesting a less disease-specific modulation. Interestingly, ocular BD showed a trend toward reduced monocyte abundance, which although not statistically significant, could reflect compartmentalization of these cells to inflamed ocular tissues, as previously described in uveitis[43, 44]. Such tissue recruitment may deplete circulating monocytes while contributing to local myeloid activation within the eye, consistent with the highly localized inflammatory pattern characteristic of ocular BD.

The heat shock proteins-enriched program we observe in non-classical monocytes from vascular BD is biologically coherent with stress-induced endothelial HSP expression acting as danger signals via TLR2/4, amplifying myeloid activation and leukocyte–endothelial interactions[45]. HSPs have been recognized as important drivers of inflammation in BD, particularly in monocytes, where they can act as an autoantigen linking microbial triggers to autoreactive T-cell responses and also induce cytokine production via TLR signaling[7]. BD patients show elevated serum of HSP60 and HSP70 families[46], and HSP60-reactive T cells are thought to contribute to vasculitis and endothelial injury[47]. This further supports a model in which HSP-driven immune activation participates in vascular inflammation and thrombotic events. Consistent with this literature, our data identify HSPA1A and HSPA1B (HSP70 family) among the top differentially expressed genes in BD monocytes, and reveal a phenotype-specific HSP70 transcriptional signature in patients with vascular BD, involving multiple HSP family members (*HSPA1A/B, HSPA8, HSPH1, HSPE1*). This vascular-skewed HSP70 program, to our knowledge not previously reported in BD, suggests that monocytes from vascular BD patients engage a heightened stress-response/chaperone network, potentially reflecting chronic exposure to inflammatory and endothelial stress signals along the vasculature.

In line with prior work demonstrating constitutive NF-κB activation and M1-skewed monocyte/macrophage polarization in BD, our ocular BD subgroup showed a classical-monocyte program heavily wired into the TNF/IL-1/NF-κB axis (**Figure 4c**). Specifically, upregulation of *NFKB1* together with the feedback inhibitor *NFKBIA* is consistent with chronic NF-κB engagement previously described in BD phagocytes and T cells, supporting persistent inflammatory signaling and apoptosis resistance [48, 49]. Concurrent overexpression of TNF superfamily members and receptors (i.e. *TNFSF8, TNFRSF10C/D, TNFRSF12A/Fn14*) suggests that classical monocytes in ocular BD are equipped to resist death signals and participate in vascular inflammation and remodeling [36, 50, 51]. The concomitant induction of *IL1B*, the IL-1 co-receptor *IL1RAP*, and the neutrophil-attracting chemokine *CXCL8* further supports an IL-1–NF-κB–neutrophil loop that mirrors the high intraocular levels of TNF-α, IL-6, and IL-8 reported in BD uveitis [52]. In light of these data, ocular BD classical monocytes might be circulating effectors of a TNF/IL-1/NF-κB–dominated program that can sustain neutrophil-rich retinal vasculitis, providing a mechanistic substrate for the previously documented efficacy of TNF-α blockade with infliximab or adalimumab in achieving rapid and sustained control of BD-associated uveitis[41, 53, 54]. Together, these results indicate that different clinical manifestations of BD may arise from distinct but overlapping myeloid activation states, emphasizing the importance of considering disease phenotypes when targeting innate immune pathways therapeutically.

Consistent with recommendations to adopt a specific phenotype approach in Behçet’s disease^1^, we analyzed activity states and vascular, ocular, and mucocutaneous phenotypes as distinct strata rather than aggregating all patients. This framework acknowledges phenotype-specific biology and is aligned with calls to study BD with due appreciation of its clinical heterogeneity, facilitating more precise mechanistic inference and phenotype-informed biomarker and therapeutic development.

This study has some limitations that should be considered when interpreting the findings. First, although we included samples from patients with active and remission BD as well as healthy controls, the sample size within each subgroup was relatively modest. This may have limited the statistical power to detect subtle transcriptional differences, particularly among less abundant immune cell subsets, and may not fully capture the biological heterogeneity of BD. In addition, because the analyses were conducted on peripheral blood mononuclear cells, the results may not fully represent the immunological alterations occurring within affected tissues, such as vascular or ocular lesions, where key pathogenic mechanisms of BD likely take place. Finally, the study is based primarily on transcriptional data, and functional validation at the protein level or through mechanistic assays will be necessary to confirm the biological relevance of the identified genes and pathways.

## 5. Conclusion

In summary, by generating a comprehensive single-cell atlas of PBMCs from patients with active and remission Behçet’s disease and healthy controls, we provide a robust transcriptomic characterization of immune cell alterations in BD. Resolving cell-type- and subset-specific transcriptional programs highlights the central role of the monocyte compartment in this vasculitis, reveals phenotype-linked myeloid states and activity-dependent plasticity, and identifies pathways likely involved in disease activity and immune dysregulation. The inclusion of clinically defined subgroups and data integration strengthens the robustness and generalizability of these findings and yields a reference resource for future studies of treatment-specific immune mechanisms in BD. Finally, the identification of reversal genes across monocyte subsets supports a model of dynamic regulatory rebalancing and suggests phenotype-aware biomarkers and therapeutic targets, motivating protein-level and tissue-based follow-up to test whether these signatures underlie clinical heterogeneity and treatment response.

## Data Availability

All data produced in the present work are contained in the manuscript or supplementary material provided

## Glossary

ADAMTS2: A Disintegrin And Metalloproteinase with Thrombospondin Motifs 2
BD: Behçet’s disease
BP: Biological Process (Gene Ontology category)
CrI: Credible Interval
DGE: Differential Gene Expression
DMARDs: Disease-Modifying Antirheumatic Drugs
DMSO: Dimethyl Sulfoxide
EDTA: Ethylenediaminetetraacetic Acid (anticoagulant)
FBS: Fetal Bovine Serum
FDR: False Discovery Rate
GBP: Guanylate-Binding Protein
GO:BP: Gene Ontology: Biological Process
GRCh38: Genome Reference Consortium human build 38 (human reference genome)
HC: Healthy Controls
HLA: Human Leukocyte Antigen
HSP: Heat-Shock Protein (e.g., HSPA1A, HSP90AA1)
HT (v4 HT kit): High-Throughput (10x Genomics Chromium Next GEM Single Cell 3′ v4 HT kit)
IFN: Interferon
IL-1β / IL-6: Interleukin-1 beta / Interleukin-6
ISG(s): Interferon-Stimulated Gene(s)
NK: Natural Killer (cell/type)
PBS: Phosphate-Buffered Saline
PBMC(s): Peripheral Blood Mononuclear Cell(s)
PCA: Principal Component Analysis
RNA-seq: RNA sequencing
scRNA-seq: Single-cell RNA sequencing
SCT / SCTransform: Seurat’s SCTransform normalization framework
SingleR: Reference-based single-cell annotation method (tool name)
TNF: Tumor Necrosis Factor
UMAP: Uniform Manifold Approximation and Projection

## 7. Acknowledgements

None

## 8. Author Contributions

E.C. contributed to experimental design, participated in sample processing, performed analyses, and wrote the manuscript. R.D. collected and processed samples and clinical data. C.B. contributed to patient recruitment and sample collection. H.D., and A.G. contributed to study design, patient recruitment and sample collection A.S. supervised the study, contributed to study design, and participated in manuscript writing. All authors discussed the results, reviewed, and approved the final version of the manuscript.

## Notes

**Conflicts of Interest:** The authors declare no conflict of interest.

### Competing Interest Statement

The authors have declared no competing interest.

### Funding Statement

This study did not receive any funding

### Author Declarations

The study was approved by the Marmara University Clinical Research Ethics Committee (Approval No: 09.2021.52).

