## Supplementary Figures for "Single-cell analysis identifies monocyte signatures of disease activity and clinical subtypes in Behçet’s disease"

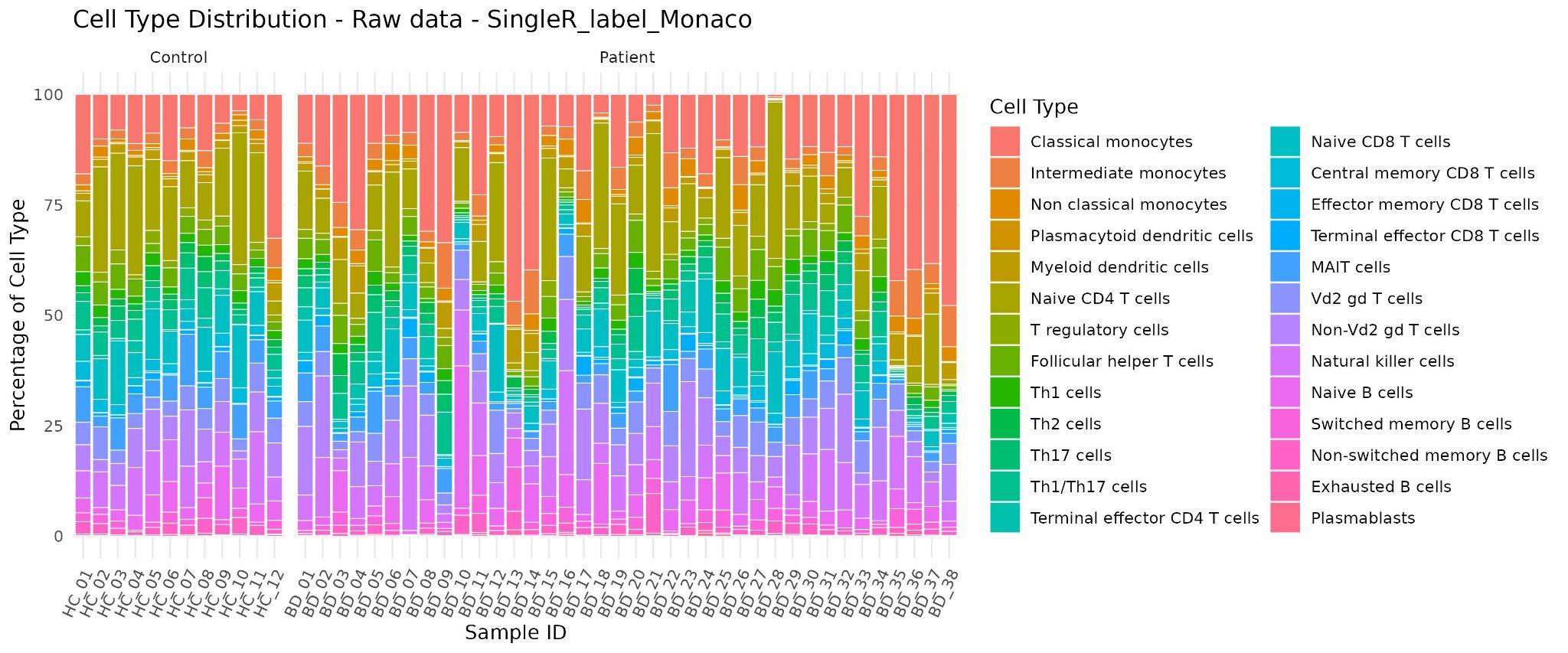
**Supplementary Figure 1. Cell type distribution across individual samples based on SingleR annotation.**

Stacked bar plots display the percentage composition of each immune cell type within peripheral blood mononuclear cells (PBMCs) for all analyzed samples, separated by healthy controls and Behçet’s disease (BD) patients. Each bar represents one sample, with colors corresponding to annotated cell populations derived from SingleR classification using the *MonacoImmuneData* reference dataset.

a

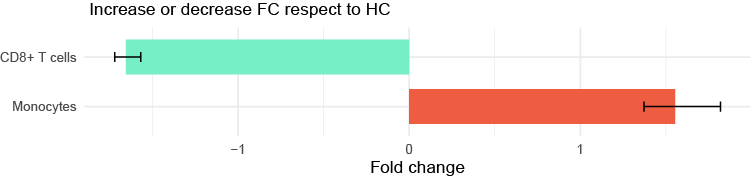

b

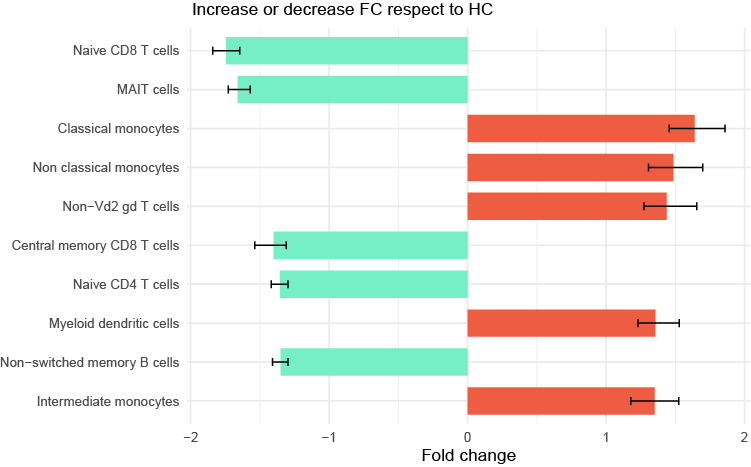

**Supplementary Figure 2. Compositional fold-change of immune cells and subsets in Behçet’s disease (BD) compared with healthy controls (HC).**
Bars represent the fold change in significant (FDR = 0.05) cell-types (a) and subsets (b) composition switches in BD relative to HC, derived from Bayesian compositional modeling with *sccomp*. Positive values denote cell types expanded in BD, while negative values indicate contraction relative to HC. Error bars indicate 95 % credible intervals for the fold-change. See Supplementary Table 3 for full statistics (effect size, credible intervals, and FDR).

a

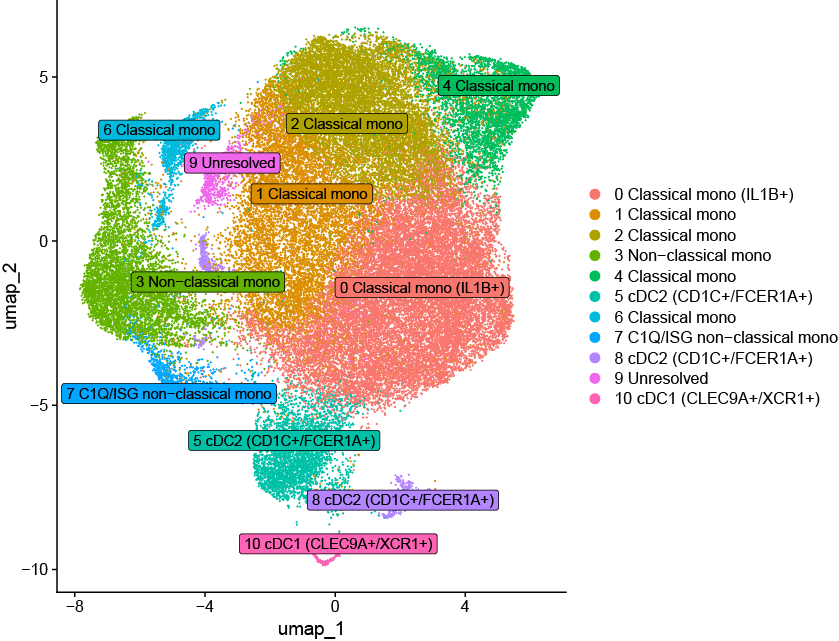

b
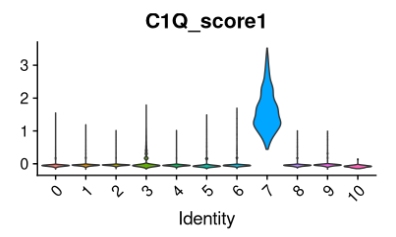

**Supplementary Figure 3.** (a) UMAP visualization of the myeloid compartment (monocytes and dendritic cells) revealing eleven transcriptionally distinct clusters, including classical, intermediate, and non-classical monocytes, as well as dendritic cell subsets (cDC1, cDC2). (b) Cluster 7 corresponds to a C1Q-enriched non-classical monocyte population, characterized by high expression of *C1QA*, *C1QB*, and *C1QC* together with interferon-stimulated genes. The violin plot shows the distribution of the C1Q module score across all clusters, confirming selective enrichment of the C1Q-high program in cluster 7.

a
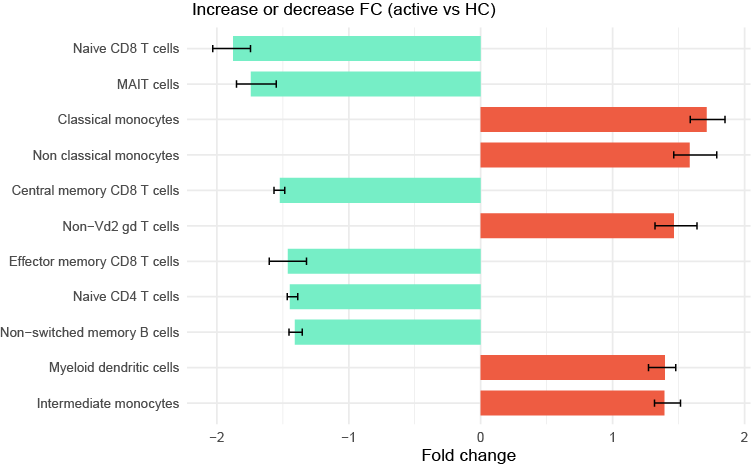

b
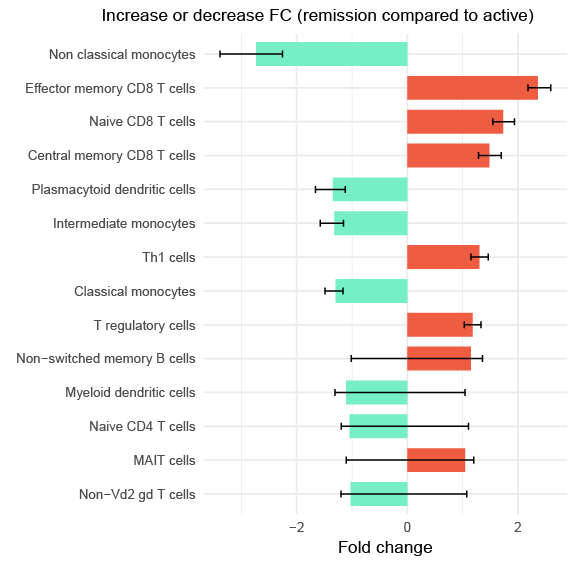

**Supplementary Figure 4. Compositional fold-change of immune cells and subsets across Behçet’s disease (BD) activity states.**Bars represent the estimated fold change in cell-type proportions obtained from Bayesian compositional modeling with sccomp. (a) Fold change comparing active BD versus healthy controls (HC), and (b) fold change comparing remission BD versus active BD. Positive values indicate cell populations expanded in the first group of each comparison, while negative values denote relative contraction. Error bars correspond to 95 % credible intervals for the fold-change estimate. See Supplementary Tables 12-13 for detailed effect sizes, and FDR-adjusted significance values.

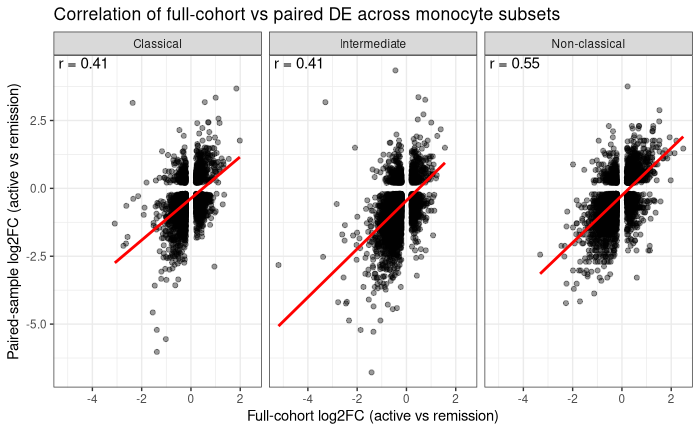

**Supplementary Figure 5. Concordance between full-cohort and longitudinal samples on differential expression analyses across monocyte subsets.**

Scatterplots show the relationship between log₂ fold-change estimates obtained from the full active-versus-remission comparison (all BD samples) and from the paired pre-/post-treatment analysis (n = 4 patient pairs) for classical, intermediate, and non-classical monocytes. Each point represents a gene tested in both analyses, with linear regression fits shown in red. Spearman correlation coefficients (r) quantify the agreement in effect sizes between the two approaches. All three subsets exhibited substantial concordance, indicating that paired samples recapitulate the major transcriptional differences observed in the full cohort despite reduced sample size.
